# Evidence For Clinical Utility of a Dimensional Measure of Autistic Traits in Toddlers

**DOI:** 10.64898/2026.10.29.26366427

**Authors:** Carolyn Lasch, Kristen Lineback, Elie Abdelnour, Holly Baer, Anna M. Abbacchi, Jennifer K. Lowe, Cheryl M. Klaiman, Stormi Lynne Pulver, Ami Klin, Daniel H. Geschwind, John N. Constantino, Natasha Marrus

**Affiliations:** Washington University in St. Louis; University of California, Los Angeles; Marcus Autism Center; Emory University School of Medicine; UCLA; Emory University

## Abstract

**Objective:** The Video-Referenced Rating of Reciprocal Social Behavior (vrRSB), a parent-report metric of quantitative autistic traits (QATs) in toddlers, leverages an innovative video scoring anchor to reliably measure heritable, continuous variation in autistic traits within the general population. However, the vrRSB’s ability to distinguish toddlers with and without autism spectrum disorder (ASD) has not been formally tested. Here we compared vrRSB scores in children with and without ASD in two samples: a community-ascertained research sample (RS) recruiting toddlers with and without ASD and a clinically referred sample (CS) consisting of toddlers with concerns for ASD.

**Methods:** T-tests and logistic regression indexed differences in vrRSB scores for toddlers with versus without ASD in both samples (RS: n_ASD_=92, n_non-ASD_=456; CS: n_ASD_=127, n_non-ASD_=44). Regression models additionally tested for differential functioning of the vrRSB relative to sex and race. ROC analysis compared classification performance of the vrRSB versus the Modified Checklist for Autism in Toddlers (M-CHAT).

**Results:** Autistic toddlers exhibited significantly higher vrRSB scores in both samples: Cohen’s *d*_RS_=3.68; Cohen’s *d*_CS_=1.04, *p*s<.001. In regression models, vrRSB scores predicted ASD diagnosis (OR=2.59, *p*<.001) and scores did not function differentially by sex or race (*p*s>.293). The vrRSB exhibited similar area under the curve as the M-CHAT in both samples and higher sensitivity in the CS.

**Conclusion:** Toddler QATs show predictive validity for autism diagnostic outcome. Consistent performance across sex and racial groups suggests promise for generalizable clinical interpretation and implementation. High sensitivity of the vrRSB supports utility for secondary screening in specialty evaluations.

**Funding statements:** Research reported in this publication was supported by the Eunice Kennedy Shriver National Institute Of Child Health & Human Development of the National Institutes of Health under Award Number P50 HD103525 to the Intellectual and Developmental Disabilities Research Center at Washington University. The content is solely the responsibility of the authors and does not necessarily represent the official views of the National Institutes of Health. This manuscript was also supported by National Institute of Mental Health (NIMH; R01MH100027 &T32 MH100019-09).

---

An ongoing challenge in the field of autism spectrum disorder (ASD or autism) is delayed early identification. Although toddler-based early intervention remains the treatment most associated with long-term improvement in functional outcomes, the median age of ASD diagnosis in the US is over 36 months^1^, after federally mandated early intervention services can be implemented. Core autism symptoms, consisting of social communication impairment and restricted repetitive interests and behaviors, can be difficult to detect in toddlers due to qualitative and developmental heterogeneity, as well as the nonlinear, accelerated growth intrinsic to typical development. This nuanced heterogeneity complicates early identification of ASD for first line providers and often requires diagnostic evaluation and confirmation by specialized clinicians with limited availability. Nevertheless, behaviors relevant to core features can inform early ASD assessment and screening, as these features, which reflect a progressive divergence from typical development^2^, can emerge prior to consolidation into ASD^3,4^.

Established screening tools for ASD (e.g., the original Modified Checklist for Autism in Toddlers (M-CHAT) and its revision, the M-CHAT-R/F^5^) have generally relied on categorical item ratings, indexing the absence of typical developmental milestones or presence of atypical behaviors that can be associated with ASD. While these tools have revolutionized general population ASD screening, reported weaknesses have included frequent false positives^6^, false negatives^7,8^ and concerns for generalizability, including in females^9^ and minority populations^10^ with historical tendencies towards reduced screening effectiveness for ASD^11^ and under-identification^12^. An alternate screening approach that may enhance early identification involves dimensional quantification of ASD-related features along a continuum, using Likert scale items rather than categorical items to rate behavior. Indexing quantitative autistic traits (QATs) in terms of levels of autistics features, rather than their presence or absence, can capture population-wide heterogeneity in ASD traits and improve sensitivity for detection of subtle or early emerging features. Dimensional measurement approaches thus have the potential to improve screening, and by extension, early identification and access to early intervention^13^ when high neuroplasticity can bolster developmental gains^14^.

From preschool-age onward, existing dimensional measures of QATs have been shown to accurately and reliably index these features. The Social Responsiveness Scale (SRS-2^15^), a well-established QAT measure used in clinical practice, can effectively screen for ASD,^16,17^ while demonstrating a continuous distribution of QATs across the general population^18,19^, indexing these traits in individuals with and without ASD. Like ASD,^20,21^ QATs indexed by the SRS are highly heritable^19,22,23^ and aggregate in families,^18,24^ with evidence supporting shared genetic factors for both ASD diagnosis and QATs^25^. In support of validity, the factor structure for the SRS aligns with DSM-5 core ASD symptom domains^26^, and, consistent with traits, QATs display developmental stability, as indicated by a test-retest correlation of 0.90-0.95 on the SRS from 8 to 12 years^27^.

Despite initial manifestations of ASD features prior to diagnosis^28^, dimensional measures of QATs have not been applied clinically to distinguish toddlers with and without ASD. The Video-Referenced Rating of Reciprocal Social Behavior (vrRSB^29^), a toddler QAT instrument, measures autistic traits in toddlers as young as 18 months, before the 30-month lower age limit of the SRS, while incorporating a video exemplar of a typically developing toddler engaged in social interaction to increase rater reliability^30^. Like the SRS, the vrRSB has demonstrated continuous, heritable variation in QATs within the general population^31^. Favorable psychometric properties include strong convergent validity with toddler measures of social competence^29^ and divergent validity with externalizing behaviors^32^. QATs on the vrRSB exhibit temporal stability with both the vrRSB itself between 18 and 24 months and with the SRS at 36 months (*r*=0.64)^32,33^ and have been preliminarily shown to distinguish toddlers with and without ASD in a small sample^30^. As a dimensional measure, the vrRSB therefore has strong potential to quantify a range of QATs in toddlers during a transitional developmental period key for enhancing screening and referral to early interventions^34,35^.

To investigate the vrRSB’s utility in distinguishing toddlers with and without ASD, we tested its performance in two samples. The first was a community-ascertained research sample (RS) consisting of toddlers recruited as typically developing or as having ASD. This sample was anticipated to capture the full range of QATs across typical and atypical development and recapitulate population-level differences in ASD traits between toddlers with and without ASD. The second sample was a real-world clinical sample (CS) of toddlers referred for a specialty clinical evaluation due to concerns for ASD. This sample comprised an at-risk population expected to show elevations in autism-related behaviors relative to the RS. Analysis focused on four questions:

1. Does the vrRSB distinguish toddlers with and without ASD in both the RS and CS?
2. Does sex affect functioning of the vrRSB?
3. Does race affect the functioning of the vrRSB?
4. How does the vrRSB perform as a screening metric, including relative to the M-CHAT, an established ASD screening tool?

We hypothesized that the vrRSB would distinguish children with and without ASD in both samples, including when considering any differential function by sex or race, based on studies of QAT measures at older ages^26^. We additionally anticipated the vrRSB, given its design as a dimensional measure focused on QATs, would explain greater variance in ASD outcome and exhibit stronger sensitivity than the M-CHAT in both samples.

## Methods

### Participants

Two toddler samples were analyzed to test the performance of the vrRSB in contexts representing differing ascertainment and levels of likelihood for ASD.

#### Research Sample (RS)

The community-recruited research sample (RS) consisted of toddler research participants identified as typically developing or as having a known or suspected ASD diagnosis. The RS pooled participants from three research studies (Table S1) conducted within a child developmental laboratory from 2013-2024 and was intended to capture a population-wide range of QATs. Participants were recruited across a Midwestern and Southern catchment area for studies characterizing early childhood manifestations of ASD and ASD traits. Toddlers in the ASD group either had a pre-existing medical diagnosis of ASD or met criteria for a DSM-based clinical best estimate diagnosis of ASD based on the Autism Diagnostic Observation Schedule (ADOS) and review of developmental assessments (Table S1). One study specifically recruited families with Black/African American heritage (referred to as Black going forward), leading to over-representation of Black participants with ASD relative to the general population. All studies were approved by the Washington University IRB.

#### Clinical Sample (CS)

The CS consisted of 17-to-30-month-old toddlers referred to a Midwestern autism clinical center between 2017-2024 due to parental or physician suspicion for ASD. It reflected a real-world, clinically ascertained population at elevated likelihood of ASD. Parents completed developmental questionnaires (including the vrRSB and M-CHAT and measures screening for delays and behavioral concerns) and presented with their toddlers for an ASD evaluation conducted by a child psychiatrist, pediatric neurologist, or nurse practitioner working under the supervision of a physician. Diagnosis was based on caregiver-report, medical chart review, observation of child play and interaction during a semi-structured play-based assessment., and questionnaires. Secondary analysis of clinical data was approved by the Washington University IRB.

### Measures

#### The Video-Referenced Rating of Reciprocal Social Behavior (vrRSB)

The vrRSB^29^ is a 44-item caregiver measure of QATs in toddlers 18-30 months old that requires 15-20 minutes to complete. QATs are indexed through Likert scale items designed to capture population-wide variation in reciprocal social behavior (RSB), with higher scores indicating greater QATs and lower RSB. The vrRSB includes 13 video-referenced items, in which informants rate a child’s RSB relative to a 3-minute video scoring anchor of a typically developing toddler providing a “frame of reference,” wherein visual context of a typically-developing toddler’s social repertoire improves both rater reliability and validity^30^. The vrRSB also includes 31 items querying examples of RSB in the last month to incorporate caregivers’ historical recall of the child’s behavior across settings. vrRSB items developmentally adapt aspects of RSB queried on the Social Responsiveness Scale (SRS-2^16^), an established QAT metric and clinical ASD screener applicable above 30 months of age. The vrRSB has demonstrated good internal and test-retest reliability^29^ and strong convergent and discriminant validity^32^. In addition to a total score, subscale scores can be calculated for the two DSM-5 based core ASD domains of social communication impairment (SCI; 33 items) and restricted, repetitive interests and behaviors (RRBs; 11 items). T-scores in analyses adjusted for age and sex (see supplemental methods for formulas) given associations previously observed in a general population sample^30^.

#### The Modified Checklist for Autism in Toddlers (M-CHAT)

The M-CHAT^36^ and Modified Checklist for Autism in Toddlers, Revised, Follow-up (M-CHAT-R/F^5^), collectively referred to as the M-CHAT, are caregiver-report questionnaires designed to screen for likelihood of ASD. These screeners contain 23 and 20 items, respectively and require 5 minutes to complete. Both have been widely used in primary care settings as Level 1 screeners for ASD, utilizing categorical yes/no questions to probe developmental progress and behaviors that tend to be delayed in children with ASD. In the RS, caregivers primarily completed the M-CHAT, while caregivers in the CS completed the M-CHAT-R, both without follow-up.

### Analytic approach

#### Descriptive Statistics

These evaluated participant characteristics by sample.

#### T-tests

T-tests evaluated vrRSB score differences in toddlers with and without ASD (Research Question 1), as well as relative to sex (Research Question 2) and race (Research Question 3) within samples and diagnostic groups. For race, Black and White populations were compared given relatively low percentages of participants among other racial groups and limited statistical power. Effect sizes are reported as Cohen’s *d*, where *d*=0.2 is small, *d*=0.5 is medium, and *d*≥0.8 is large.

#### Logistic regressions

A series of regressions quantified how changes in vrRSB t-score affected likelihood of an ASD diagnosis (Research Question 1), and whether this varied by participant sex (Research Question 2) or participant race (Research Question 3). Initial models examined effects of sample ascertainment, including an interaction term for sample x vrRSB. The addition of sex and an interaction term for sex x vrRSB then tested whether the vrRSB’s relationship to ASD outcome differed in males and females; a significant interaction term or significantly better model performance indicated differential function of the vrRSB by sex. An analogous series of models was extended to White and Black participants to explore potential differential functioning of the vrRSB by race.

To compare performance of the vrRSB and M-CHAT (Research Question 4), logistic regressions were fit separately in the RS and CS. Models included: 1) a baseline model with participant sex (Model 1); 2) an M-CHAT pass/fail variable added to Model 1 (Model 2); 3) a vrRSB pass/fail variable added to Model 1 (Model 3); and 4) a model with both M-CHAT and vrRSB pass/fail variables to assess unique contributions to variance by measure (Model 4). Pass/fail variables were used to conform to clinical screening practices. For the M-CHAT, scores based on the original M-CHAT were transformed to a ‘1’ in accordance with guidelines^36^ in which either 2 positively scored critical items or 3 or more positively scored items in total indicated a positive screen. For the M-CHAT-R/F, the published high-risk categorization with 40% or more items endorsed^5^ was classified as a ‘1,’ while scores below this threshold were transformed to ‘0’. For the vrRSB, the pass/fail t-scores cut-off was ≥ 65 (i.e., 1.5 SD above the population mean of 50), modeled on the screening cut-off applied in the SRS-2^16^ (see Figure S1 for ROC curve of vrRSB t-scores in both samples).

Model fit across regressions was compared using the corrected Akaike Information Criterion (AICc^37^), Bayes Information Criterion (BIC^38^) and R^2^ values. AICc provides an estimation of model fit compared to other models in a family built in the same sample. BIC is more rewarding of parsimony, and more readily ‘punishes’ overfitting of models in small samples. Differences in AICc and BIC of 2 or more represent positive evidence of significant differences in model fit^39,40^. R^2^ values evaluated differences in variance explained across models.

#### Calculation of psychometric performance

In addition to model comparisons of ASD predictive validity, the area under the curve (AUC), sensitivity, specificity, accuracy, positive predictive value (PPV), and negative predictive value (NPV), (see Table S2 for formulas), were compared for the vrRSB and M-CHAT in the RS and CS (Research Question 4). Psychometric performance measures were also calculated for the SCI and RRB subscales of the vrRSB (Table S6). R packages pROC^41^ and epiR^42^ calculated psychometric measures and confidence intervals. Significant differences between performance metrics were informally evaluated based on non-overlapping confidence intervals.

## Results

### Participant Characteristics

The research sample (RS) included 552 participants (47% female) with a mean age of 23.8 +/- 2.5 months (Table 1). ASD prevalence was 17% (9% and 25% in females and males, respectively). Most RS participants endorsed being either White (68%) or Black (20%), although the percentage of Black toddlers among the ASD group was relatively high (82% in Black toddlers versus 7% in White toddlers) due to one study’s focused recruitment of Black children with ASD. Toddlers with versus without ASD showed a higher percentage positive M-CHAT screens, 73% versus 9%, (χ^2^(1)=166.70, p<.001).

**Table 1.** Participant demographics.

| Measures | Recruitment and Diagnostic Group |  |  |  |
|---|---|---|---|---|
|  | Research Sample |  | Clinical Sample |  |
|  | ASD | No ASD | ASD | No ASD |
| N | 96 | 456 | 127 | 44 |
| Age; months (SD) | 25.0 (3.2) | 23.6 (2.2) | 23.7 (3.2) | 23.4 (3.4) |
| Range | 18 - 29 | 16 - 29 | 18 - 29 | 16 - 29 |
| Race |  |  |  |  |
| White | 14 (15%) | 364 (80%) | 73 (58%) | 31 (71%) |
| Black/African American | 79 (82%) | 33 (7%) | 45 (35%) | 8 (18%) |
| Asian | 3 (3%) | 1 (<1%) | 3 (2%) | 3 (7%) |
| Pacific Islander | -- | -- | 2 (2%) | -- |
| More than one reported | 6 (6%) | 64 (14%) | -- | -- |
| Unknown or not reported | -- | 10 (2%) | 4 (3%) | 2 (5%) |
| Ethnicity |  |  |  |  |
| Endorsed Hispanic | 1 (1%) | 206 (45%) | -- | 1 (2%) |
| Sex |  |  |  |  |
| Male | 72 (75.0%) | 222 (48.7%) | 96 (75.6%) | 31 (70.5%) |
| Female | 24 (25.0%) | 234 (51.3%) | 31 (24.4%) | 13 (29.5%) |
| vrRSB Total |  |  |  |  |
| t-score mean (SD) | 100.0 (23.1) | 49.1 (10.9) | 99.3 (23.8) | 75.2 (21.6) |
| Range | 45 - 151 | 27 - 111 | 42 - 166 | 42 - 135 |
| M-CHAT (n) | 79 | 352 | 104 | 33 |
| Above follow-up threshold N (%) | 57 (73%) | 30 (9%) | 69 (66%) | 7 (21%) |

Note: Participants endorsing more than one race are counted in all races endorsed and in ‘multiracial’. Thus, sum of racial categories does not sum to 100%. Ethnicity not reported in the Clinical Sample, as this information was not recorded for clinical care.
Note: ASD=autism spectrum disorder, vrRSB=Video-Referenced Rating of Reciprocal Social Behavior; M-CHAT=Modified Checklist of Autism in Toddlers

**Figure 1.**
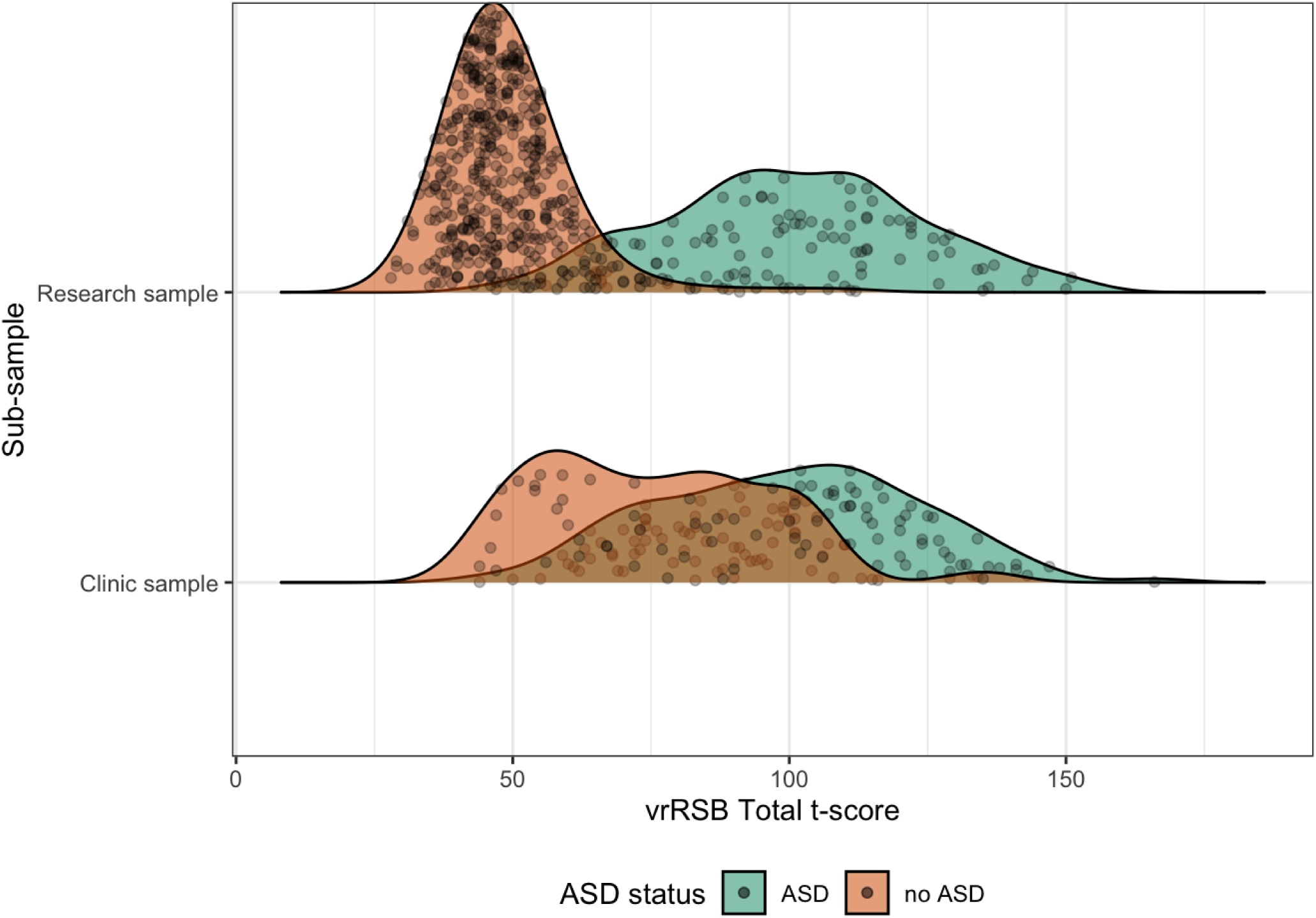
Distributions of vrRSB scores (sex- and age-normed) as a factor of participant sex, recruitment source, and ASD diagnosis.

The clinic sample (CS) included 171 participants (26% female) with a mean age of 23.6 +/- 3.2 months and ASD prevalence of 74% (71% and 76% in females and males, respectively), greater than the RS. Like the RS, most participants endorsed being either White (61%) or Black (31%), with a similar breakdown among toddlers with ASD. The percentage of positive M-CHAT screens was higher in toddlers with ASD versus without ASD, 66% versus 21%, (χ^2^(1)=62.46, p<.001). A larger percentage of toddlers without ASD screened positive compared to the RS (21% versus 9%), consistent with enrichment of the CS for ASD-related concerns.

### vrRSB Score Differences Relative to ASD Diagnosis

vrRSB t-scores showed substantial differences between autistic and non-autistic toddlers in both samples. In the RS, a large effect, *d*=3.68, 95% CI [3.37, 3.99], characterized mean score differences for toddlers with versus without ASD: M_ASD_=100.1 versus M_noASD_ =49.1, *t*(104)=21.1, *p*<.001. In the elevated-likelihood CS, toddlers diagnosed with ASD also showed higher vrRSB t-scores, M_ASD_=99.3, than toddlers not diagnosed with ASD, M_noASD_=75.1, *t*(82)=6.22, *p*<.001, also with a large effect size of *d*=1.04, 95% CI [0.68, 1.40].

Comparisons of vrRSB scores for toddlers with and without ASD (Table S4, Figure S2) showed similar effect sizes for both males and females in the RS (*ds*≥3.13) and the CS (*ds*≥0.74). vrRSB scores for males and females with and without ASD strongly overlapped (Figure S3) and no sex differences were found in toddlers with ASD. Among CS participants without ASD, vrRSB t-scores were nominally higher for females (M=84.9) than males (M=71.1), *p=*0.057. vrRSB t-scores unadjusted for sex showed similar findings (Table S4), except for slightly higher scores in males versus females in the RS for toddlers without ASD (*d*=0.36, 95% CI [0.17, 0.54]).

Similarly, large differences in vrRSB t-scores (Table S5; Figure S4) were observed between toddlers with and without ASD for both Black and White participants in the RS (*ds*≥2.18) and CS (*ds*≥0.94). For both the RS and CS, no significant score differences were observed between Black and White participants with ASD *(ps*>0.486) or without ASD (*p*s>0.216, Figure S5).

### Models of vrRSB Associations with ASD

A series of logistic regression models (Table 2) examined the vrRSB’s ability to predict ASD diagnostic outcome and whether this varied by recruitment sample, sex, or race. In the baseline Model 1, main effects of recruitment sample (RS vs. CS), vrRSB t-score, and their interaction were all highly significant. Participants in the elevated-likelihood CS, which exhibited higher ASD prevalence, were more likely to receive an ASD diagnosis than those in the RS (OR=7.29, *p*<0.001). Higher vrRSB t-scores were also associated with increased likelihood of ASD and displayed a significant interaction with sample type (*p*<0.001). For the RS, each additional point increased the odds of ASD by 13.7%, while this effect was only 4.7% in the CS, which reflected a higher overall likelihood of ASD.

**Table 2.**
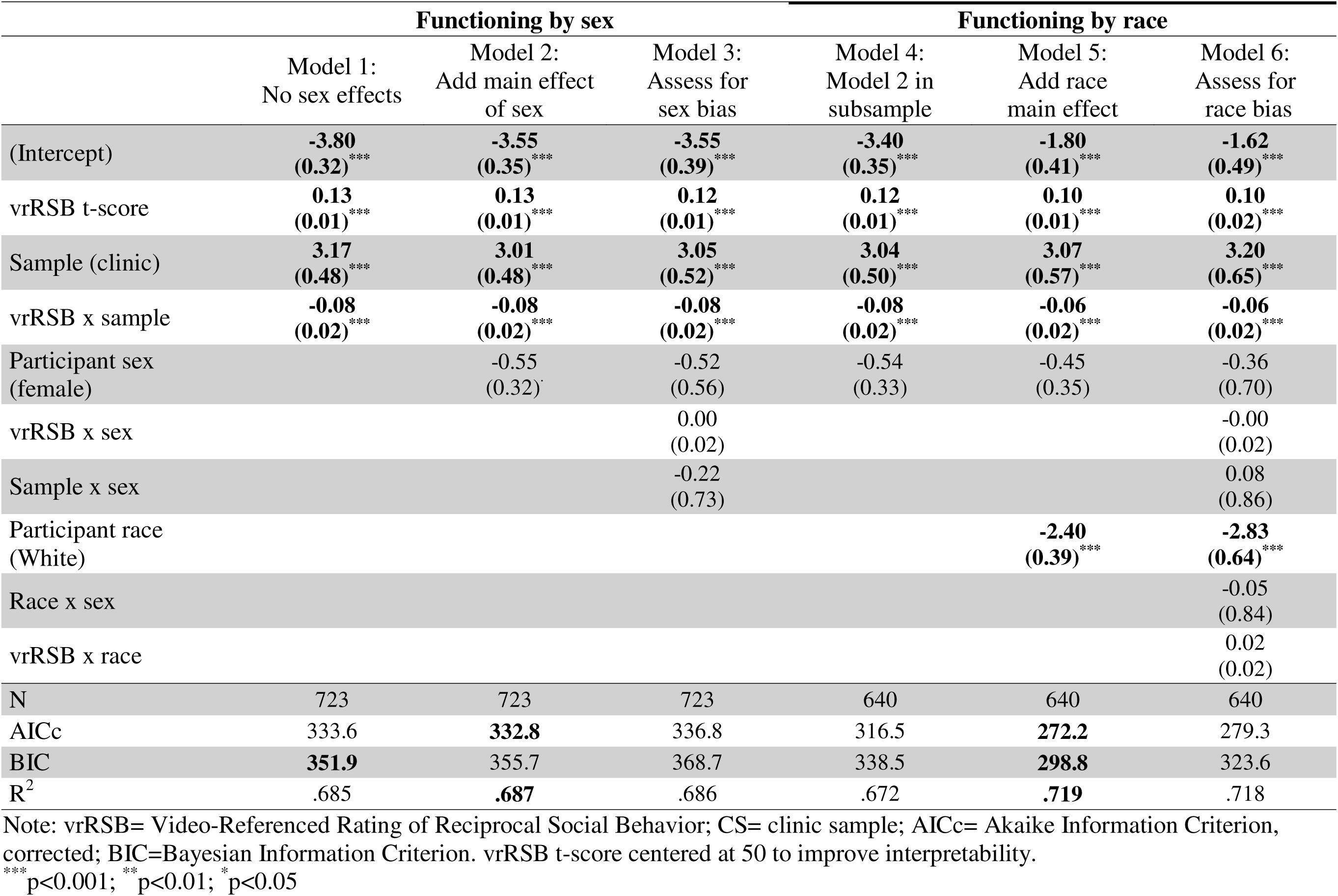
vrRSB logistic models predicting ASD and examining effects of sex and race on vrRSB performance.

|  | Functioning by sex |  |  | Functioning by race |  |  |
|---|---|---|---|---|---|---|
|  | Model 1:<br>No sex effects | Model 2:<br>Add main effect of sex | Model 3:<br>Assess for sex bias | Model 4:<br>Model 2 in subsample | Model 5:<br>Add race main effect | Model 6:<br>Assess for race bias |
| (Intercept) | **-3.80**<br>**(0.32)**<sup>\*\*\*</sup> | **-3.55**<br>**(0.35)**<sup>\*\*\*</sup> | **-3.55**<br>**(0.39)**<sup>\*\*\*</sup> | **-3.40**<br>**(0.35)**<sup>\*\*\*</sup> | **-1.80**<br>**(0.41)**<sup>\*\*\*</sup> | **-1.62**<br>**(0.49)**<sup>\*\*\*</sup> |
| vrRSB t-score | **0.13**<br>**(0.01)**<sup>\*\*\*</sup> | **0.13**<br>**(0.01)**<sup>\*\*\*</sup> | **0.12**<br>**(0.01)**<sup>\*\*\*</sup> | **0.12**<br>**(0.01)**<sup>\*\*\*</sup> | **0.10**<br>**(0.01)**<sup>\*\*\*</sup> | **0.10**<br>**(0.02)**<sup>\*\*\*</sup> |
| Sample (clinic) | **3.17**<br>**(0.48)**<sup>\*\*\*</sup> | **3.01**<br>**(0.48)**<sup>\*\*\*</sup> | **3.05**<br>**(0.52)**<sup>\*\*\*</sup> | **3.04**<br>**(0.50)**<sup>\*\*\*</sup> | **3.07**<br>**(0.57)**<sup>\*\*\*</sup> | **3.20**<br>**(0.65)**<sup>\*\*\*</sup> |
| vrRSB x sample | **-0.08**<br>**(0.02)**<sup>\*\*\*</sup> | **-0.08**<br>**(0.02)**<sup>\*\*\*</sup> | **-0.08**<br>**(0.02)**<sup>\*\*\*</sup> | **-0.08**<br>**(0.02)**<sup>\*\*\*</sup> | **-0.06**<br>**(0.02)**<sup>\*\*\*</sup> | **-0.06**<br>**(0.02)**<sup>\*\*\*</sup> |
| Participant sex (female) |  | -0.55<br>(0.32)<sup>.</sup> | -0.52<br>(0.56) | -0.54<br>(0.33) | -0.45<br>(0.35) | -0.36<br>(0.70) |
| vrRSB x sex |  |  | 0.00<br>(0.02) |  |  | -0.00<br>(0.02) |
| Sample x sex |  |  | -0.22<br>(0.73) |  |  | 0.08<br>(0.86) |
| Participant race (White) |  |  |  |  | **-2.40**<br>**(0.39)**<sup>\*\*\*</sup> | **-2.83**<br>**(0.64)**<sup>\*\*\*</sup> |
| Race x sex |  |  |  |  |  | -0.05<br>(0.84) |
| vrRSB x race |  |  |  |  |  | 0.02<br>(0.02) |
| N | 723 | 723 | 723 | 640 | 640 | 640 |
| AICc | 333.6 | **332.8** | 336.8 | 316.5 | **272.2** | 279.3 |
| BIC | **351.9** | 355.7 | 368.7 | 338.5 | **298.8** | 323.6 |
| R<sup>2</sup> | .685 | **.687** | .686 | .672 | **.719** | .718 |

Note: vrRSB= Video-Referenced Rating of Reciprocal Social Behavior; CS= clinic sample; AICc= Akaike Information Criterion, corrected; BIC=Bayesian Information Criterion. vrRSB t-score centered at 50 to improve interpretability.
<sup>\*\*\*</sup>p<0.001; <sup>\*\*</sup>p<0.01; <sup>\*</sup>p<0.05

### Testing for Differential vrRSB Function by Sex and Race

In Model 2, an additional main effect of participant sex was not significant and did not substantively improve model fit (AICc decrease <2). In Model 3, neither an interaction term of the vrRSB with sex (*p=*0.75) nor sample with sex (*p*=0.81) improved model fit. In both models, the effects of vrRSB score, sample, and their interaction remained highly significant (*p*s<.001). Model 1 provided the most parsimonious fit based on the lowest BIC. Thus, the vrRSB did not evidence differential functioning by participant sex.

For evaluating differential functioning of the vrRSB by race, a baseline model (Model 4) included vrRSB score, sample, sex, and an interaction term for score and sample; and showed significant effects for all terms except sex. An additional main effect term for race in Model 5 showed greater odds of ASD in Black participants (*p*<0.001), aligning with enriched recruitment of Black toddlers with autism in the RS. In contrast, an interaction term for race x vrRSB (Model 6) was not significant (*p*=0.29) and did not improve model fit, indicating that the vrRSB does not function differentially for White vs. Black participants.

### Comparison of ASD association for vrRSB and M-CHAT

Logistic regressions evaluated the performance of the vrRSB and M-CHAT separately in the RS (n=431) and CS (n=137) for participants with data on both measures. In the RS (Table 3), a model including only the vrRSB outperformed a model with only the M-CHAT, 67.6% versus 39.8% of variance explained. The overall best-fitting model included both measures (69.0% of variance explained). In the CS, the vrRSB also outperformed the M-CHAT (23.1% vs. 15.9% of variance explained) and the best-fitting model included both measures (26.2% of variance explained).

**Table 3.** Models predicting ASD diagnosis in the RS and CS with vrRSB and M-CHAT.

|  | Research Sample (RS; N=431) |  |  |  | Clinic Sample (CS; N=137) |  |  |  |
|---|---|---|---|---|---|---|---|---|
|  | Baseline | M-CHAT | vrRSB | M-CHAT + vrRSB | Baseline | M-CHAT | vrRSB | M-CHAT + vrRSB |
| (Intercept) | **-1.04<br>(0.15)<sup>\*\*\*</sup>** | **-2.36<br>(0.25)<sup>\*\*\*</sup>** | **-4.22<br>(0.54)<sup>\*\*\*</sup>** | **-4.33<br>(0.55)<sup>\*\*\*</sup>** | **1.26<br>(0.24)<sup>\*\*\*</sup>** | 0.44<br>(0.28) | -0.59<br>(0.44) | -0.65<br>(0.44) |
| Participant sex (female) | **-1.22<br>(0.28)<sup>\*\*\*</sup>** | **-0.87<br>(0.34)<sup>\*</sup>** | -0.39<br>(0.48) | -0.39<br>(0.49) | -0.43<br>(0.45) | -0.67<br>(0.50) | -0.96<br>(0.51)<sup>.</sup> | -1.02<br>(0.54)<sup>.</sup> |
| M-CHAT |  | **3.28<br>(0.32)<sup>\*\*\*</sup>** |  | **1.07<br>(0.48)<sup>\*</sup>** |  | **2.06<br>(0.48)<sup>\*\*\*</sup>** |  | **1.39<br>(0.54)<sup>\*\*</sup>** |
| vrRSB |  |  | **5.56<br>(0.56)<sup>\*\*\*</sup>** | **4.96<br>(0.60)<sup>\*\*\*</sup>** |  |  | **2.64<br>(0.54)<sup>\*\*\*</sup>** | **1.95<br>(0.58)<sup>\*\*\*</sup>** |
| AIC | 393.81 | 270.33 | 152.56 | **149.70** | 154.38 | 134.18 | 128.87 | **123.98** |
| BIC | 401.94 | 282.53 | **164.75** | 165.97 | 160.22 | 142.94 | 137.63 | **135.66** |
| R<sup>2</sup> | 0.046 | 0.398 | 0.676 | **0.690** | 0.007 | 0.159 | 0.231 | **0.262** |

Note: M-CHAT= Modified Checklist of Autism in Toddlers; vrRSB=Video-Referenced Rating of Reciprocal Social Behavior; CS=clinic sample; AICc=Akaike Information Criterion, corrected; BIC=Bayesian Information Criterion
Note: vrRSB uses a cutoff at t-score of 65. M-CHAT uses a cutoff of a raw score of 4 on the M-CHAT and a raw score of 8 on the M-CHAT-R.
<sup>\*\*\*</sup>p<0.001; <sup>\*\*</sup>p<0.01; <sup>\*</sup>p<0.05

### Screening Performance Metrics for the vrRSB and M-CHAT

In the RS, the vrRSB out-performed the M-CHAT on several metrics (Table 4). AUC for the vrRSB was strong, 0.943, and confidence intervals did not overlap with AUC_M-CHAT_=0.825, reflecting a significant difference. The vrRSB’s accuracy, sensitivity, and NPV were also strong and significantly greater than the M-CHAT, with a particularly large difference for sensitivity, which was 0.949 for the vrRSB and 0.734 for the M-CHAT. The vrRSB thus detected 29% more ASD cases than the M-CHAT, and the M-CHAT did not identify any additional positive cases missed by the vrRSB (Table S6).

**Table 4.** Psychometric performance of the vrRSB and M-CHAT.

| Screening measure & sample | ASD- | ASD+ | AUC | Specificity | Sensitivity | Accuracy | PPV | NPV |
|---|---|---|---|---|---|---|---|---|
| Research (RS) |  |  |  |  |  |  |  |  |
| vrRSB 65-t-score cutoff |  |  |  |  |  |  |  |  |
| Screen ASD- | 330 | 4 | **0.943\***<br>[0.916, 0.971] | 0.938<br>[0.907, 0.960] | **0.949\***<br>[0.875, 0.986] | **0.940\***<br>[0.913, 0.960] | 0.773<br>[0.677, 0.852] | **0.988\***<br>[0.970, 0.997] |
| Screen ASD+ | 22 | 75 |  |  |  |  |  |  |
| M-CHAT cutoff |  |  |  |  |  |  |  |  |
| Screen ASD- | 322 | 21 | 0.825<br>[0.773, 0.876] | 0.915<br>[0.881, 0.942] | 0.734<br>[0.623, 0.827] | 0.882<br>[0.847, 0.911] | 0.659<br>[0.550, 0.757] | 0.939<br>[0.908, 0.962] |
| Screen ASD+ | 30 | 58 |  |  |  |  |  |  |
| Clinic (CS) |  |  |  |  |  |  |  |  |
| vrRSB 65-t-score cutoff |  |  |  |  |  |  |  |  |
| Screen ASD- | 16 | 8 | 0.704<br>[0.614, 0.794] | 0.485<br>[0.308, 0.665] | **0.923\***<br>[0.854, 0.966] | 0.818<br>[0.743, 0.878] | 0.850<br>[0.770, 0.910] | 0.667<br>[0.447, 0.844] |
| Screen ASD+ | 17 | 96 |  |  |  |  |  |  |
| M-CHAT cutoff |  |  |  |  |  |  |  |  |
| Screen ASD- | 26 | 35 | 0.726<br>[0.641, 0.810] | 0.788<br>[0.611, 0.910] | 0.663<br>[0.564, 0.753] | 0.663<br>[0.609, 0.769] | 0.908<br>[0.819, 0.962] | 0.426<br>[0.300, 0.559] |
| Screen ASD+ | 7 | 69 |  |  |  |  |  |  |

Note: M-CHAT= Modified Checklist of Autism in Toddlers; vrRSB= Video-Referenced Rating of Reciprocal Social Behavior; AUC= Area under the curve, PPV= positive predictive value, NPV= negative predictive value
Note: Bolded with\*=significantly better performance than the other measure (non-overlapping CIs)
Note: M-CHAT uses a cutoff of a raw score of 4 on the M-CHAT and a raw score of 8 on the M-CHAT-R.

In the CS, AUC values were good for both measures and nominally higher for the M-CHAT (AUC_vrRSB_=0.704, AUC_M-CHAT_=0.726), given overlapping confidence intervals. M-CHAT metrics were also nominally higher for PPV (0.908 versus 0.850 for the vrRSB) and specificity (0.788 versus 0.485). As with the RS, the vrRSB showed significantly higher sensitivity, 0.923 versus 0.663 for the M-CHAT, identifying 39% more ASD cases than the M-CHAT. As in the CS, the M-CHAT did not identify any cases missed by the vrRSB (Table S6).

Comparison of performance metrics for vrRSB SCI and RRB subscales relative to total scores showed nominally higher performance metrics for total scores in both samples (Table S6). The SCI subscale showed significantly higher sensitivity than the RRB subscale (0.962 versus 0.788) in the CS and nominally higher sensitivity in the RS (0.977 versus 0.861). The RRB subscale showed significantly higher specificity than the SCI subscale in the CS (0.545 versus 0.212) and nominally higher specificity in the RS (0.895 vs. 0.777).

## Discussion

Results strongly support utility of the vrRSB as a clinically relevant tool for enhancing early diagnosis of ASD. As hypothesized, the vrRSB displayed large effect sizes differentiating toddlers with and without ASD, including in a real-world clinical population with elevated concern for ASD. Psychometric strengths included the lack of differential functioning by sex and race and several comparable or better screening performance metrics than the M-CHAT, an established ASD screener. These strengths suggest that a dimensional approach to ASD screening, informed by etiologically relevant, heritable features of ASD, can enhance early identification, with implications for accelerating access to early intervention.

Large vrRSB score differences between toddlers with and without ASD in the RS, *d*=3.68, correspond to the clinically established SRS, for which QATs are on average 3 SD higher in individuals with ASD versus the general population^16^. This magnitude is especially notable for toddlers, considering non-trivial assessment challenges in this age group due to variability in developing social capacities and active ASD symptom consolidation. A large effect size was observed in the at-risk CS sample as well, *d*=1.04, highlighting the robustness of this measure in a more ambiguous context. These results support utility of the vrRSB as an adjunctive measurement tool in real-world specialty clinical evaluation, often an essential step for early ASD diagnosis. Additionally, items referencing the scoring anchor of a typically-developing toddler’s social repertoire help reduce interpretation discrepancies, increasing reliability, while the dimensional nature of the items allows raters to identify nuance in a child’s abilities and behavior.

The vrRSB did not display differential function by race or sex, as scores behaved similarly across males and females and Black and White participants. Score ranges across these groups consistently overlapped. Effect sizes distinguishing toddlers with and without ASD were large and did not vary significantly based on participant sex or racial background. Further, interaction terms testing for differential function in regressions were not significant. Sex differences in mean scores for specific diagnostic subgroups were modest and agreed with known properties of ASD traits. For example, higher scores in males in the non-ASD group for the RS (Table S4) were consistent with greater ASD traits in males previously observed in the general population^25^, while numerically higher scores in females without ASD in the CS aligned with evidence for a higher ASD liability threshold in females, i.e., a female protective effect^43^. Because both sex and race have been associated with reduced screening effectiveness, as well as under-diagnosis and misdiagnoses^12,30^, metrics that function similarly across demographic groups are important for improving equity in screening and avoiding diagnostic and treatment delays^44,45^ during a vital developmental window for optimizing outcomes.

Performance metrics for diagnostic group discrimination by the vrRSB also demonstrated multiple strengths. In the RS, which represented a population-wide range of ASD traits, AUC, accuracy, specificity, and sensitivity were excellent, while in the elevated-likelihood CS, sensitivity remained excellent. Collectively, the high sensitivity in both samples, as well as a high NPV in the RS, underscore the vrRSB’s ability to minimize false negatives, similar to the SRS at older ages. This property is especially advantageous for higher-prevalence clinical contexts with toddler populations, when avoiding missed diagnoses is a priority to avoid children missing critical opportunities for early intervention. Clinically, the vrRSB could potentially serve a unique role as a) an adjunctive assessment tool to enhance diagnostic confidence in high-risk clinical populations or b) a screen to assist with triaging evaluations in overburdened specialty centers, which could accelerate identification in referred toddlers most likely to have ASD.

In comparing performance of the vrRSB and M-CHAT, the vrRSB consistently explained more variance, including more unique variance, in line with its specific focus on autistic traits. In the RS, which represented population-wide variation in ASD traits, vrRSB performance metrics generally met or exceeded the M-CHAT. Further, higher sensitivity of the vrRSB in both samples showcased its value-added ability to detect a substantial percentage of ASD cases (>28%) missed by M-CHAT thresholds. Although the M-CHAT’s specificity was greater than the vrRSB in the CS, the M-CHAT did not uniquely identify any toddlers with ASD in either sample. The lower specificity of the vrRSB specifically in the CS may reflect a psychometric trade-off relative to the dimensional approach of quantifying heritable ASD traits. Human genetic studies have demonstrated shared genetic liability across psychiatric conditions^46^, consistent with findings of elevated autistic traits in school-aged children with other developmental concerns (e.g., ADHD) who do not meet ASD criteria^47^. Thus, the enhanced biological relevance of a QATs, by invoking transdiagnostic relationships, may reduce discriminatory capacity, particularly in clinical populations more likely to feature co-occurring conditions. Given the relative strength in specificity noted for RRB items, adding RRB items could be considered to enhance vrRSB screening performance.

Our findings are limited by the cross-sectional nature of the ASD diagnostic data relative to diagnostic stability. Prospective studies have observed that some toddlers meet ASD diagnostic criteria after 30 months while other toddlers may no longer meet diagnostic criteria later in childhood^48^, despite generally high diagnostic stability after age 24 months^49^. Thus, performance metrics could vary with long-term follow-up. We were also unable to account for other possible causes of differential functioning across both samples, such as socioeconomic status (SES), caregiver education, or English fluency. Future larger, more diverse samples would also allow broader examination of differential functioning relative to other demographic factors, including at the level of individual items, and inform optimization of the vrRSB.

While the stronger performance of the vrRSB relative to the M-CHAT in the RS could stem from differences in RS and CS toddler composition, it could also reflect greater use of the M-CHAT versus M-CHAT-R in the RS. The RS pooled across several studies to maximize statistical power, but this then included data collected prior to broad dissemination of the M-CHAT-R. Additionally, in the CS, clinicians were able to reference both the M-CHAT and vrRSB during diagnostic assessment, which could have inflated measurement performance, highlighting the need to test for replication of findings in studies separating screening measures from evaluation of ASD diagnosis.

As a more practical limitation, the vrRSB requires about 15 minutes to complete, whereas the M-CHAT and many first-line screeners take 5 minutes or less. Leveraging its psychometric advantages clinically may therefore be best suited to staged screening or to inform Level 2 evaluation of children at risk of ASD. Nevertheless, as a dimensional trait measure reflecting heritable ASD liability across the population while also indexing early ASD diagnostic outcome, the vrRSB may serve dual clinical and biologically informative research applications. The stability^32,33^ of the vrRSB during the period when ASD features arise and consolidate could allow characterization of developmental mechanisms, as well as neural and genetic correlates of ASD and ASD liability that could refine future risk assessment or more personalized targeting of treatment. The vrRSB could also be applied as part of longitudinal surveillance to monitor consolidation of autistic traits relative to hypothesized protective or risk factors, or used to test hypothesized predictive associations between infant temperament and ASD trait outcomes^50^ that can be measured population-wide, rather than limited to only children who develop ASD.

## Conclusion

Analyses support the use of the vrRSB for effectively identifying autistic toddlers in both population-wide contexts and clinical settings. QATs in toddlers represented variance related to ASD outcomes, and the vrRSB’s psychometric performance metrics were comparable to or better than the M-CHAT, a widely used screener. The vrRSB’s psychometric strengths suggest it could be especially helpful to avoid missing ASD cases in clinical evaluation of high-risk populations, or triaging children for more in-depth evaluation following initial screening. While further validation testing is warranted, strategic clinical application of the vrRSB in clinical practice could enhance early identification and clinical efficiency, promoting more timely delivery of early intervention.

## Data Availability

All data produced in the present study are available upon reasonable request to the authors.

