## Supplemental Information for "Evidence For Clinical Utility of a Dimensional Measure of Autistic Traits in Toddlers"

**Supplementary Information**

#### Supplemental Methods

#### Participants

**Recruitment of RS participants**. Recruitment of participants in the Research Sample included use of fliers, world of mouth, and social media postings such as parent groups/ pages on sites such as Facebook.

Table S1. Information on sub-samples combined to form Research Sample

| **Study Name** | **Population Type** | **N** | **Age Range (mos.)** | **N with ASD** | **How ASD was assessed** |
| --- | --- | --- | --- | --- | --- |
| Early Reciprocal Social Behavior (ERSB)^1^ | General Population  Epidemiologically sampled twin pairs (one twin per pair)  Some have ASD, some community recruited | 354 | 16-30 | 3 | - Community-based Diagnosis or positive ADOS-2 - vrRSB at 18 mos. - Preschool SRS-2 at 36 months |
| Genetics of Neurodevelopment in African Americans (GENAA)^2^ | Black/African American  ASD, suspected ASD, and control | 89 | 18-30 | 79 | - ADOS-2 - Cognitive (Mullen, Raven’s, DAS, PPVT-5) - Vineland - DSM interview - SRS-2 and SCQ |
| Social Motivation Project | Community-recruited | 109 | 16-29 | 14 | - Community diagnosis |

Note: ASD=autism spectrum disorder; Dx =diagnosis; ADOS=Autism Diagnostic Observation Schedule, 2^nd^ edition; vrRSB= video-referenced rating of Reciprocal Social behavior; SRS-2=Social Responsiveness Scale, 2^nd^ edition; DAS=Differential Abilities Scale; PPVT-5= Peabody Picture Vocabulary Test, 5^th^ edition; DSM= Diagnostic Statistical Manual; SCQ=Social Communication Questionnaire.

Note: participants in the GENAA study did not exclusively endorse Black/ African American backgrounds and thus included individuals who also reported Hispanic ethnicity (1%) and / or multiple racial groups (9%).

#### Supplemental measures information

**T-score creation for the vrRSB**. The vrRSB scores were transformed from raw to t-score in R using the following steps:

1. vrRSB_predicted_=33.77041-(0.56446*(age months))-(2.48926*female)
2. vrRSB_z-score_=(vrRSB_raw_ – vrRSB_predicted_)/ 8.994983
3. vrRSB t-score=floor( 50+ (10* vrRSB_z-score_)

Formulas were derived using samples described in Marrus et al. (2018) *JOVE* paper. As seen from the formula in step 1, on average raw vrRSB total sum scores for males were 2.5 raw points higher than females.

#### Supplemental Analytic Approach information

Performance of vrRSB without sex-control of t-**scores**.

T-tests evaluating sex differences within subgroups of the research sample (RS) and clinical sample (CS) were repeated using a non-sex-controlled vrRSB t-score to evaluate whether sex differences in raw scores were present, given prior studies showing sex differences in vrRSB scores not adjusted for sex (e.g., Marrus et al, 2015).

##### Definitions of measure performance metrics.

Table S2. Measures of measure performance used to compare vrRSB and M-CHAT.

| Term | Definition | Formula |
| --- | --- | --- |
| AUC | Average value of specificity for all possible sensitivity values |  |
| Accuracy | Probability of being categorized correctly | (TN+TP)/ total |
| Sensitivity | Probability of a positive test if you have ASD. | TP/ (TP+FN) |
| Specificity | Probability of a negative test if you do not have ASD | TN/ (TN+FP) |
| Positive predictive value (PPV) | Likelihood that a person with a positive test result is a True Positive | TP/ (TP+ FP) |
| Negative predictive value (NPV) | The likelihood that a person who has a negative test result is a True Negative | TN/ (TN +FN) |

*Figure S1.* ROC curves for vrRSB Total t-score in the Clinic Sample (CS, black) and Research Sample (RS, red). Large circles indicate the location of t-score of 65 on each curve.
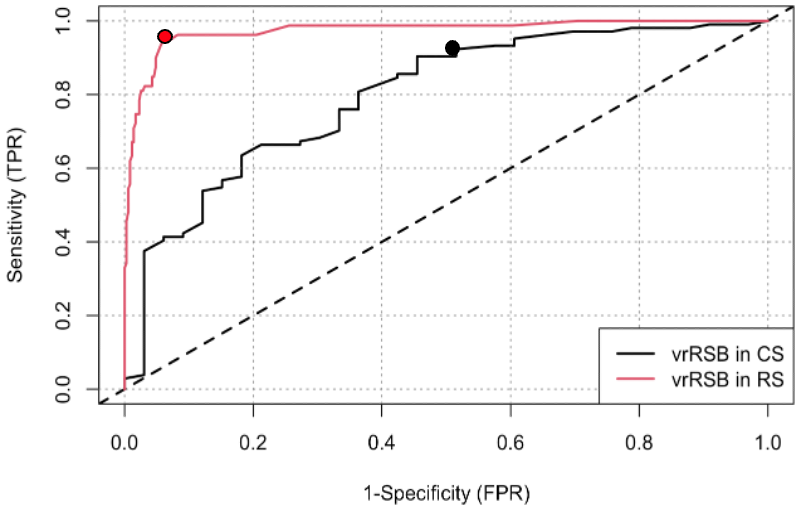

ROC curves were modeled with R package ROCit^3^.

***Comparison of vrRSB and M-CHAT with continuous predictors***. Additional regressions were run using continuous scores on the vrRSB (age- and sex-controlled t-scores used in rest of main analyses) and percent of items endorsed on M-CHAT. As with the main analyses, regressions were carried out separately in community and CS samples, and measures of performance were also computed for each sample using continuous scores.

### Supplemental Results

#### Supplemental participant information

Table S3. Participant demographics by diagnosis, recruitment sample, and in full combined sample.

| Measures | Recruitment or Diagnostic Group | | | | |
| --- | --- | --- | --- | --- | --- |
|  | ASD | No ASD | Research Sample (RS) | Clinical Sample (CS) | Full Sample |
| N | 223 | 500 | 552 | 171 | 723 |
| Age; months (SD) | 24.2 (3.24) | 23.5 (2.33) | 23.8 (2.46) | 23.6 (3.23) | 23.8 (2.66) |
| Range | 18 - 29 | 16 - 29 | 16 - 29 | 16 - 29 | 16-29 |
| Race |  |  |  |  |  |
| White | 87 (39%) | 395 (79%) | 378 (68%) | 104 (61%) | 482 (67%) |
| Black/African American | 124 (56%) | 41 (8%) | 112 (20%) | 53 (31%) | 165 (23%) |
| Asian | 6 (3%) | 4 (<1%) | 4 (<1%) | 6 (4%) | 10 (1%) |
| Pacific Islander | 2 (<1%) | -- | -- | 2 (1%) | 2 (<1%) |
| More than one reported | 6 (3%) | 64 (13%) | 70 (13%) | -- | 70 (10%) |
| Unknown or not reported | 4 (2%) | 12 (2%) | 10 (2%) | 6 (4%) | 16 (2%) |
| Ethnicity |  |  |  |  |  |
| Endorsed Hispanic | 1 (<1%) | 207 (41%) | 207 (38%) | 1 (<1%) | 208 (29%) |
| Sex |  |  |  |  |  |
| Male | 168 (75%) | 253 (51%) | 294 (53%) | 127 (74%) | 421 (58%) |
| Female | 55 (25%) | 247 (49%) | 258 (47%) | 44 (26%) | 302 (42%) |
| vrRSB Total |  |  |  |  |  |
| t-score mean (SD) | 99.6 (23.4) | 51.4 (14.3) | 58.0 (23.7) | 93.1 (25.5) | 66.3 (28.4) |
| Range | 42-166 | 27-135 | 27-151 | 42-166 | 27-166 |
| M-CHAT (n) | 183 | 385 | 431 | 137 | 568 |
| Above follow-up threshold (%) | 125 (68%) | 10 (3%) | 61 (14%) | 74 (54%) | 135 (24%) |
| Note: Participants endorsing more than one race are counted in all races endorsed and in ‘More than one reported’. Thus, count of racial categories does not sum to 100%.  Note: Ethnicity missing in Clinical Sample, as this information was not recorded for clinical care.  ASD=autism spectrum disorder, vrRSB=video-referenced scale of Reciprocal Social Behavior; M-CHAT=Modified Checklist for Autism in Toddlers (combines original and Revised/ Follow-up versions) | | | | | |

#### vrRSB differentiates ASD, with and without sex-normed scores

Table S4. Comparison of effect of ASD on vrRSB t-scores when controlling for sex in vrRSB scoring (column 3) vs. no sex-control (column 4)

| T-test comparison | Subsample (N) | Cohen’s *d* with  sex-norming of t-scores | Cohen’s *d* without  sex-norming of t-scores |
| --- | --- | --- | --- |
| ASD 🡪 vrRSB | CS (171) | 1.04  95% CI [0.68, 1.40]  ASD=99.3, noASD=75.2 | 1.05  95% CI [0.69, 1.41]  ASD=99.5, noASD =75.5 |
| ASD 🡪 vrRSB | CS males (127) | 1.18  95% CI [0.75, 1.62]  ASD=98.6, noASD=71.1 | 1.18  95% CI [0.75, 1.62]  ASD=99.4, noASD=72.2 |
| ASD 🡪 vrRSB | CS females (44) | 0.74  95%CI [0.05, 1.420  ASD=101.6, noASD=84.9 | 0.73  95% CI [0.05, 142]  ASD=99.7, noASD=83.2 |
| ASD 🡪 vrRSB | RS (552) | 3.68  95% CI [3.37, 3.99]  ASD=100.0, noASD=49.1 | 3.70  95% CI [3.39, 4.01]  ASD=100.2, noASD=49.0 |
| ASD 🡪 vrRSB | RS males (294) | 3.13  95% CI [2.76, 3.50]  ASD=99.8, noASD=49.7 | 3.14  95% CI [2.77, 3.51]  ASD=100.6, noASD=51.0 |
| ASD 🡪 vrRSB | RS females (258) | 4.79  95% CI [4.20, 5.38]  ASD=100.8, noASD=48.5 | 4.77  95% CI [4.18, 5.36]  ASD=98.9, noASD=47.1 |
| Sex 🡪 vrRSB | CS ASD (127) | -0.13  95% CI [-0.54, -.28]  M=98.6, F=101.6 | -0.01  95% CI [-0.42, 0.40]  M=99.4, F=99.7 |
| Sex 🡪 vrRSB | RS ASD (96) | -0.04  95% CI [-0.51, 0.43]  M=99.8, F=100.8 | 0.07  95% CI [-0.34, 0.54]  M=100.6, F=98.9 |
| Sex 🡪 vrRSB | CS no ASD (44) | -0.66  95% CI [-0.02, 1.35]  M=73.6, F=88.7 | -0.54  95% CI [-0.14, 1.21]  M=72.2, F=83.2 |
| Sex 🡪 vrRSB | RS no ASD (127) | 0.11  95% CI [-0.07, 0.30]  M=49.7, F=48.5 | 0.36  95% CI [ 0.17, 0.54]  M=51.0, F=47.1 |

Note: ASD= autism spectrum disorder, vrRSB= Video-Referenced Rating of Reciprocal Social Behavior, RS= research sample, CS=clinic sample

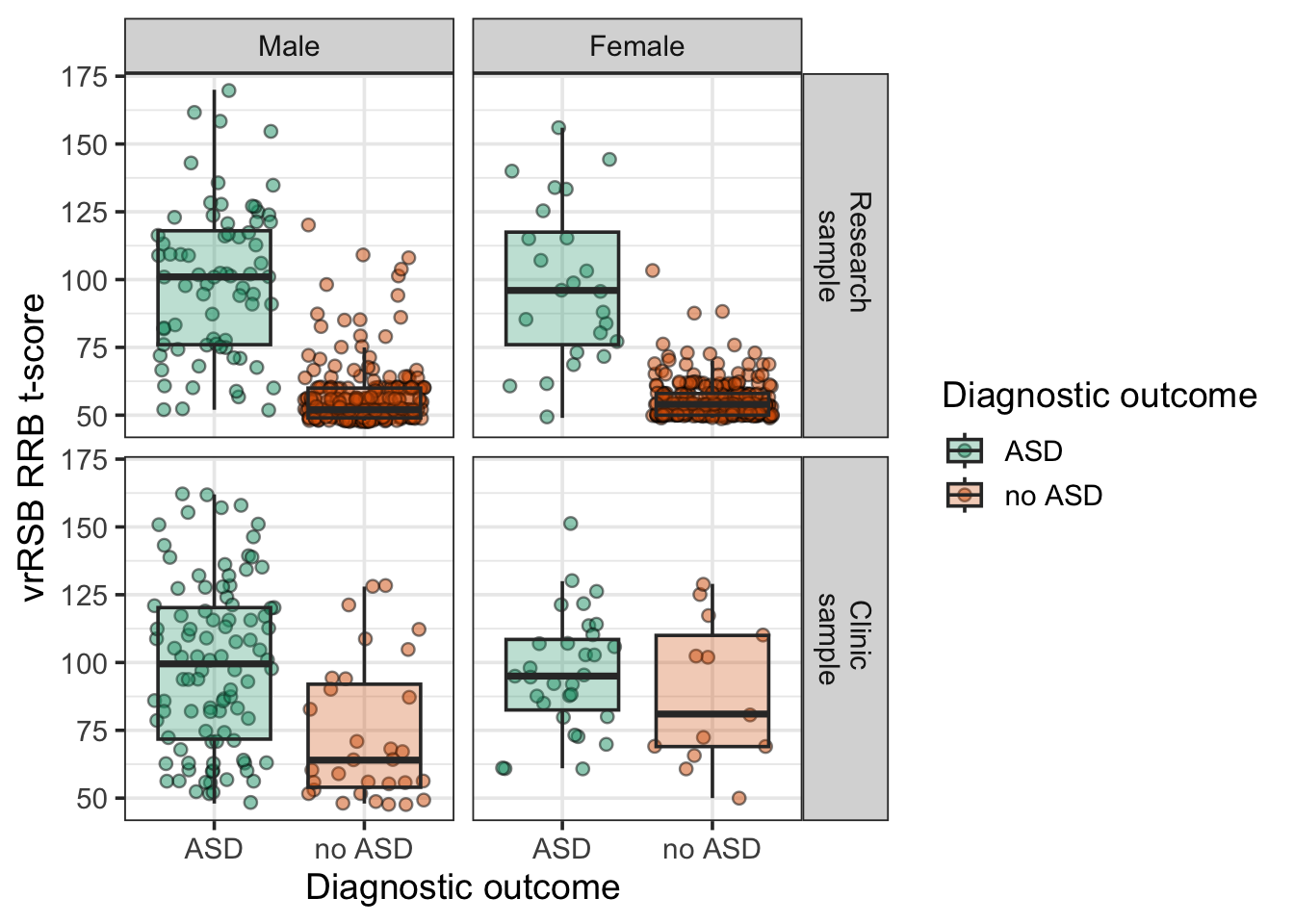

*Figure S2*. vrRSB clearly differentiates between ASD (green) and non-ASD (orange) participants in research sample (top) and clinic sample (bottom), as well as in males (left) and females (right).

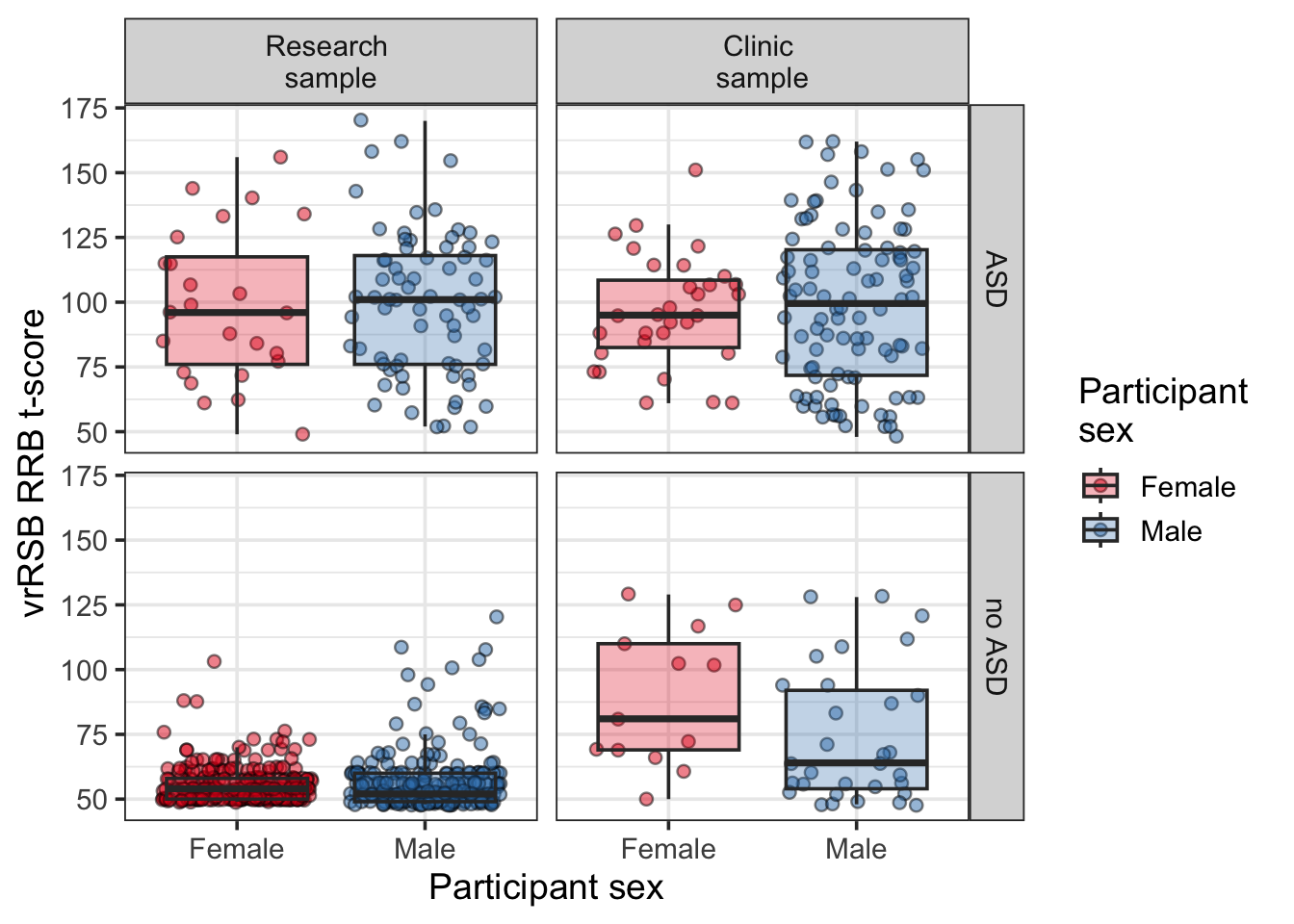

*Figure S3*. vrRSB scores do not differ for males (blue) and females (red) with ASD (top), nor for non-autistic participants from the research sample (bottom left). Non-autistic females in the clinic sample have marginally higher vrRSB t-scores than their male counterparts (bottom right).

Table S5. Comparison of effect of ASD on vrRSB t-scores in Black/ AA vs. non-Black/AA subsamples, and effects of participant racial background on vrRSB t-scores by recruitment and diagnostic subsample.

| T-test comparison | Subsample (N) | Cohen’s *d* of t-scores |
| --- | --- | --- |
| ASD 🡪 vrRSB | RS Black (112) | 2.18  95% CI [1.68, 2.68]  ASD=100.0, noASD=52.7 |
| ASD 🡪 vrRSB | RS White (440) | 4.25  95% CI [3.57, 4.92]  ASD=95.5, noASD=49.0 |
| ASD 🡪 vrRSB | CS Black (53) | 0.97  95% CI [0.17, 1.76]  ASD =97.9, noASD=75.0 |
| ASD 🡪 vrRSB | CS White (118) | 0.94  95% CI [0.50, 1.39]  ASD=100.2, noASD=78.2 |
| Racial background 🡪 vrRSB | RS ASD (96) | 0.19  95% CI [-0.45, 0.83]  Black = 100.0, White=95.5 |
| Racial background 🡪 vrRSB | RS no ASD (456) | 0.33  95% CI [-0.03, 0.69]  Black = 52.7, White=49.0 |
| Racial background 🡪 vrRSB | CS ASD (127) | -0.09  95% CI [-0.47, 0.28]  Black = 97.9, White = 100.2 |
| Racial background 🡪 vrRSB | CS no ASD (44) | -0.15  95% CI [-0.95, 0.66]  Black= 75.0, White = 78.2 |

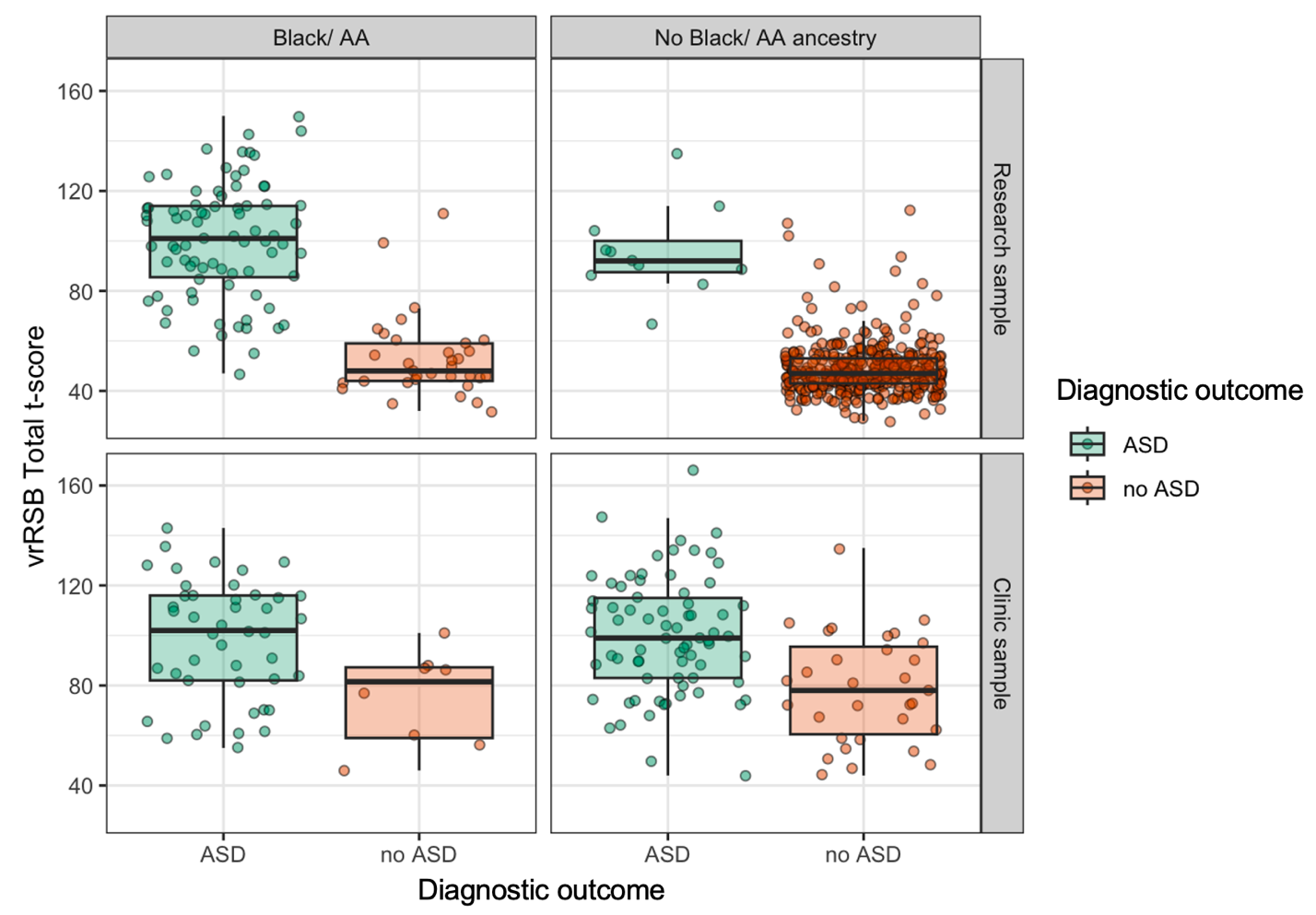
*Figure S4*. vrRSB clearly differentiates between ASD (green) and non-ASD (orange) participants in research sample (top) and clinic sample (bottom), as well as in participants with Black/ African American ancestry (left) and those without (right).

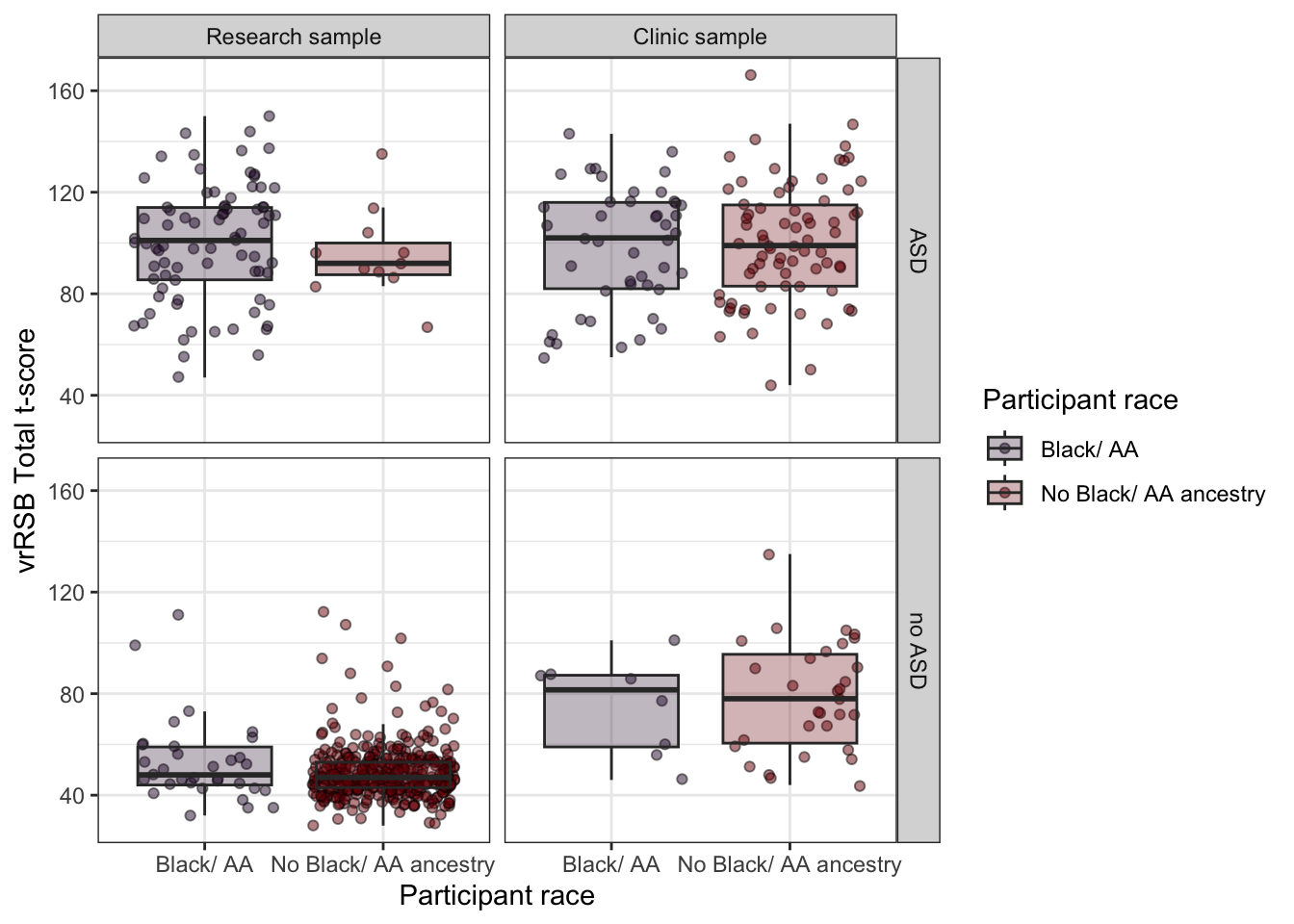

*Figure S5*. vrRSB scores do not differ for participants with Black/ African American ancestry (purple) and those without (red). This is true in participants with ASD (top) and for non-autistic participants (bottom). It is also true in the Research Sample (left) and the Clinic Sample (right).

Table S5. *ASD True Positives and True Negatives Uniquely Detected by vrRSB and M-CHAT.*

|  | Unique true positives identified | | Unique true negatives identified | |
| --- | --- | --- | --- | --- |
|  | RS | CS | RS | CS |
| vrRSB | 17 | 27 | 19 | 1 |
| MCHAT | 0 | 0 | 11 | 11 |

*Note.* ASD=autism spectrum disorder, vrRSB=Video-Referenced Rating of Reciprocal Social Behavior; M-CHAT=Modified Checklist of Autism in Toddlers; RS= research sample; CS= clinic sample.

*Note*: In the RS, total ASD cases were 79, and total non-ASD cases were 372. In the CS, total ASD cases were 104, and total non-ASD cases were 33.

#### Comparison of vrRSB SCI and RRB subscales in comparison to full vrRSB

Psychometric performance variables were calculated for the vrRSB SCI and RRB subscales to compare their performance to the full vrRSB scale in research and clinical context. To binarize continuous scores to approximate a cut off approach to screening, t-score of 65 and above on Total, SCI, and RRB vrRSB scales were binarized to ‘1’, and t-score below 64 were binarized to 0.

In both samples, vrRSB Total score had better performance than either subscale, such that estimated AUC, specificity, PPV, etc. was highest for the total score, but with 95% confidence intervals that overlapped with those of the SCI and RRB subscales.

- RS
  - Total has sig better AUC, specificity, accuracy, PPV than SCI
  - SCI has nominally better sensitivity and NPV than total and RRB .
  - Total has nominally better PPV Than RRB
  - **RRB has significantly better accuracy than SCI** (but nominally lower than total)
- CS
  - Total nominally better AUC, accuracy than SCI and RRB
  - **RRB nominally better specificity than total** (which is nominally better than SCI)
  - SCI has significantly better sensitivity than RRB, and nominally better than total

Table S6. *Statistical performance of vrRSB subscales (SCI and RRB) compared to vrRSB Total t-score*

| Screening Measure + sample | AUC | Specificity | Sensitivity | Accuracy | PPV | NPV |
| --- | --- | --- | --- | --- | --- | --- |
| Research sample |  |  |  |  |  |  |
| vrRSB 65-t-score cutoff | .943  (.916- .971) | .938  (.907- .960) | .949  (.875- .986) | .940  (.913- .960) | .773  (.677- .852) | .988  (.970- .997) |
| SCI 65 t-score cutoff | .868  (.840- .896) | .777  (.729- .820) | .977  (.919 - .997) | .817  (.777 - .852) | 0.522  (0.442- 0.601) | 0.993  (0.973- 0.999) |
| RRB 65 t-score cutoff | .878  (836-.920) | 0.895  (0.858- 0.925) | 0.861  (0.765- 0.928) | ***0.889***  (0.855- 0.917) | 0.648  (0.548- 0.738) | 0.966  (0.940 - 0.983) |
| Clinic sample |  |  |  |  |  |  |
| vrRSB 65-t-score cutoff | .704  (.614-.794) | .485  (.308-.665) | **.923***  (.854-.966) | .818  (.743-.878) | .850  (.770-.910) | .667  (.447-.844) |
| SCI 65 t-score cutoff | .587  (.514 - .660) | 0.212  (0.090- 0.389) | ***0.962***  (0.904- 0.989) | 0.781  (0.702- 0.847) | 0.794  (0.712- 0.861) | 0.636  (0.308- 0.891) |
| RRB 65 t-score cutoff | .667  (.572-.762) | 0.545  (0.364- 0.719) | 0.788  (0.697- 0.862) | 0.730  (0.647- 0.802) | 0.845  (0.758- 0.911) | 0.450  (0.293- 0.615) |

Note: **Bolded with*=**significantly better performance than the other measure (non-overlapping CIs)

Note: AUC=Area under the curve, PPV=positive predictive value, NPV=negative predictive value; vrRSB= Video-Referenced Rating of Reciprocal Social Behavior; SCI= Social communication impairment; RRB=restricted, repetitive behaviors; M-CHAT=Modified Checklist for Autism in Toddler
